# Detection of Tuberculosis by The Analysis of Exhaled Breath Particles with High-resolution Mass Spectrometry

**DOI:** 10.1101/2020.04.20.20073163

**Authors:** Dapeng Chen, Wayne A. Bryden, Robin Wood

## Abstract

Tuberculosis remains a global health threat killing over 1 million people *per* year. Current sputum-based diagnostics are specific but lack sensitivity resulting in treatment of many sputum negative cases. In this proof-of-concept study, we used high-resolution mass spectrometry to identify specific lipids in peripheral lung fluid samples of TB patients and controls, captured using a novel non-invasive sampling system. Exhaled respiratory particles were collected in liquid and after concentration and lipid extraction directly infused into a high-resolution mass spectrometer. High-resolution mass spectrometric data collection was conducted in a dual ion mode and chemical compositions were constructed using accurate mass measurement. Over 400 features with high segregating capacity were extracted and optimized using feature selection algorithm and machine learning, from which the accuracy of detection of positive tuberculosis patients was estimated. This current strategy provides sensitivity offered by high-resolution mass spectrometry and can be readily susceptible for developing a novel clinical assay exploring peripheral lung fluid for the detection of active TB cases.

## Introduction

Tuberculosis (TB) is caused by *Mycobacterium tuberculosis* and contributed to 1.6 million deaths worldwide in 2017^1^. It is accepted that the detection of active TB cases is critical to initiate treatment and stop ongoing transmission^2^. However, 40% of TB incident cases were under-detected due to the lack of rapid detection methods in the most TB-affected areas^2^. Generally, TB cases are only reported to the local healthcare community by patients when severe symptoms of pulmonary TB occur^2^. Molecular technology-based detection methods offer several advantages comparing to conventional sputum microscopy and several diagnostic technologies relying on nucleic acid amplification (polymerase chain reaction, PCR) were endorsed by WTO as advanced diagnostic tests for TB^3,4^. Although PCR-based diagnostic tools exhibit high sensitivity and have the capacity to detect multiple drug-resistant TB cases, the requirement of using sputum and lengthy turnaround time limit its application for rapid screening in high TB-burden communities^3^. Therefore, there is a great need for the development of new screening tests that are operationally feasible in high-burdened settings and can rapidly and accurately detect active TB disease at a low cost-per-test. The promising application of mass spectrometry (MS) gains attention in the fields of environmental measurements, health care, and clinical diagnostics, etc^5-7^. Introduction of high-resolution orbitrap mass spectrometers (>100, 000 FWHM, full-width-at-half-maximum) enables the unambiguous assignment of chemical compositions without performing tandem mass spectrometry experiments^8,9^. Conventionally, high-resolution mass spectra can be collected within a few minutes, which provides ultra-high mass accuracy and low false positive rate for molecular composition assignment^9^. For this reason, it offers a potential for development of an advanced diagnostic tool with improved sensitivity, enhanced dynamic range, and unparalleled mass accuracy^10^.

In previous studies, we developed a breath aerosol sampling unit, Respiratory Aerosol Sampling Chamber (RASC), to collect exhaled bioaerosol particles emitted from the lungs of active TB patients^11,12^. In this study, we developed a strategy to exploit the bioaerosol sampling system and high-resolution mass spectrometry for the characterization of biomolecules for the detection of positive TB cases.

## Results

### Extraction method validation using two lipid standards

Sample introduction to mass spectrometry requires small injection volume. For this reason, we optimized an extraction method to reduce the sample complexity and concentrate breath samples by targeting lipid species in breath samples. Two commercial standard lipids, 1,2-Distearoyl-sn-glycero-3-phosphorylcholine (C_44_H_88_NO_8_P) and 1,2-Dilauroyl-sn-glycero-3-phosphorylcholine (C_32_H_64_NO_8_P), were used as quality control to validate lipid extraction methods and mass spectrometry performance in this study (Fig. 1). Mass spectrometry analysis showed that the full masses of two lipid standards are 622.4440 and 790.6340 (MH+, Fig. 1 A) and the identification of an identical diagnostic fragment ion of phosphorylcholine (m/z 184.0724, Figs. 1, B and C, Fig. S1) was acquired by higher-energy collision dissociation (HCD) with 35% of energy. The extraction efficiency was evaluated by using the extracted signal intensity of two lipid standards and the results showed that extraction efficiency is slightly higher than 50%.

**Figure 1.**
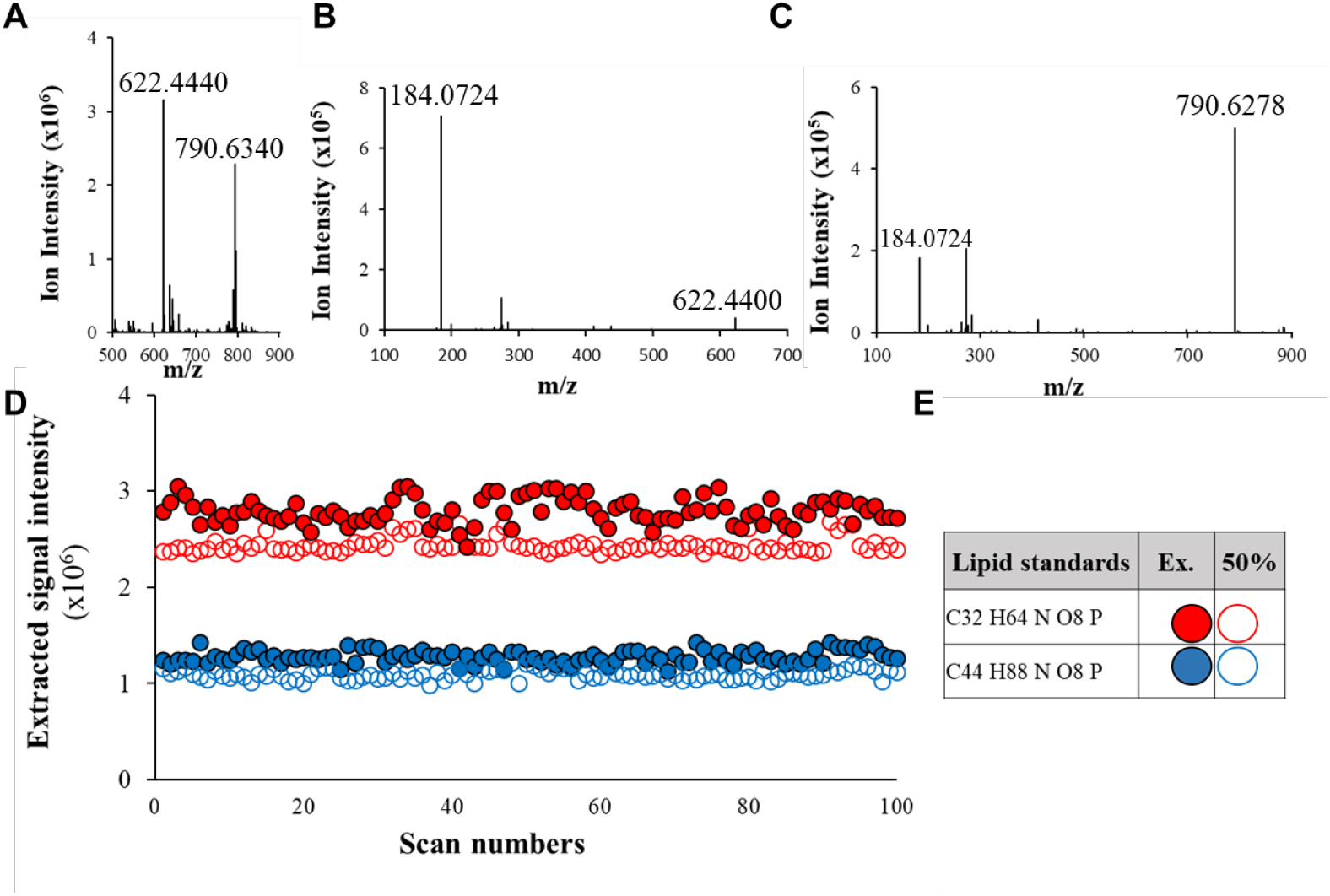
Evaluation of lipid extraction method using lipid standards and highresolution mass spectrometry. High-resolution mass spectrum of two lipid standards **(A)**, 1,2-Distearoyl-sn-glycero-3-phosphorylcholine (C_44_H_88_NO_8_P) and 1,2-Dilauroyl-sn-glycero-3-phosphorylcholine (C_32_H_64_NO_8_P), and the fragment ion patterns **(B, C)**. The extraction efficiency was evaluated by the comparison of the signal intensity of two lipid standards **(D)**. Hallow red and blue dots represent the signal intensity of 50% of the starting material of two lipid standards used for the method validation. Solid red and blue dots represent signal intensity of lipid standards after extraction **(E)**.

### High-resolution mass spectrometry data collection on non-Tb subjects and TB patients

Data collection was conducted in the mass range of 200 to 2000 m/z in positive ion mode (Fig. 2 A) and 400 to 2000 m/z in negative ion mode (Fig. 2 B). It is immediately apparent that non-TB and TB samples show different mass spectrometric patterns. TB samples generated strong signals in the mass range of 400 to 1400 m/z in the positive ion mode and 1100 to 2000 m/z in the negative ion mode (Fig. 2 A). PCA was applied to features extracted from both ion modes with a signal to noise ratio cutoff (S/N>5, Figs. 2 C-E). Over 4000 features were extracted from positive ion mode and over 2500 features from negative ion mode. PCA results showed that non-TB subjects were prone to cluster together and there was a separation trend between two classes of samples using features extracted from positive ion mode in the mass range from 200 to 2000 m/z (Fig. 2 C). However, the analysis of four TB patient samples overlapped with non-TB subject samples (Fig. 2 C). In negative ion mode, PCA was initially applied to the features collected from the mass range from 400 to 2000 m/z (Fig. 2 D). The results showed that samples from either class were prone to congregate and a poor separation was observed (Fig. 2 D), suggesting over-loading of features in PCA. Data collection in negative ion mode shows that non-TB and TB mass spectrometric profiles had distinguishable patterns in the mass range of 800 to 2000 m/z. Therefore, to achieve a better separation in the negative ion mode, features extracted from the mass range of 900 to 2000 m/z were performed and better segregation was observed between these two classes of samples (Fig. 2 E). In addition, this more defined mass range in negative ion mode resulted in a reduction in the number of features from ∼ 2500 to 763.

**Figure 2.**
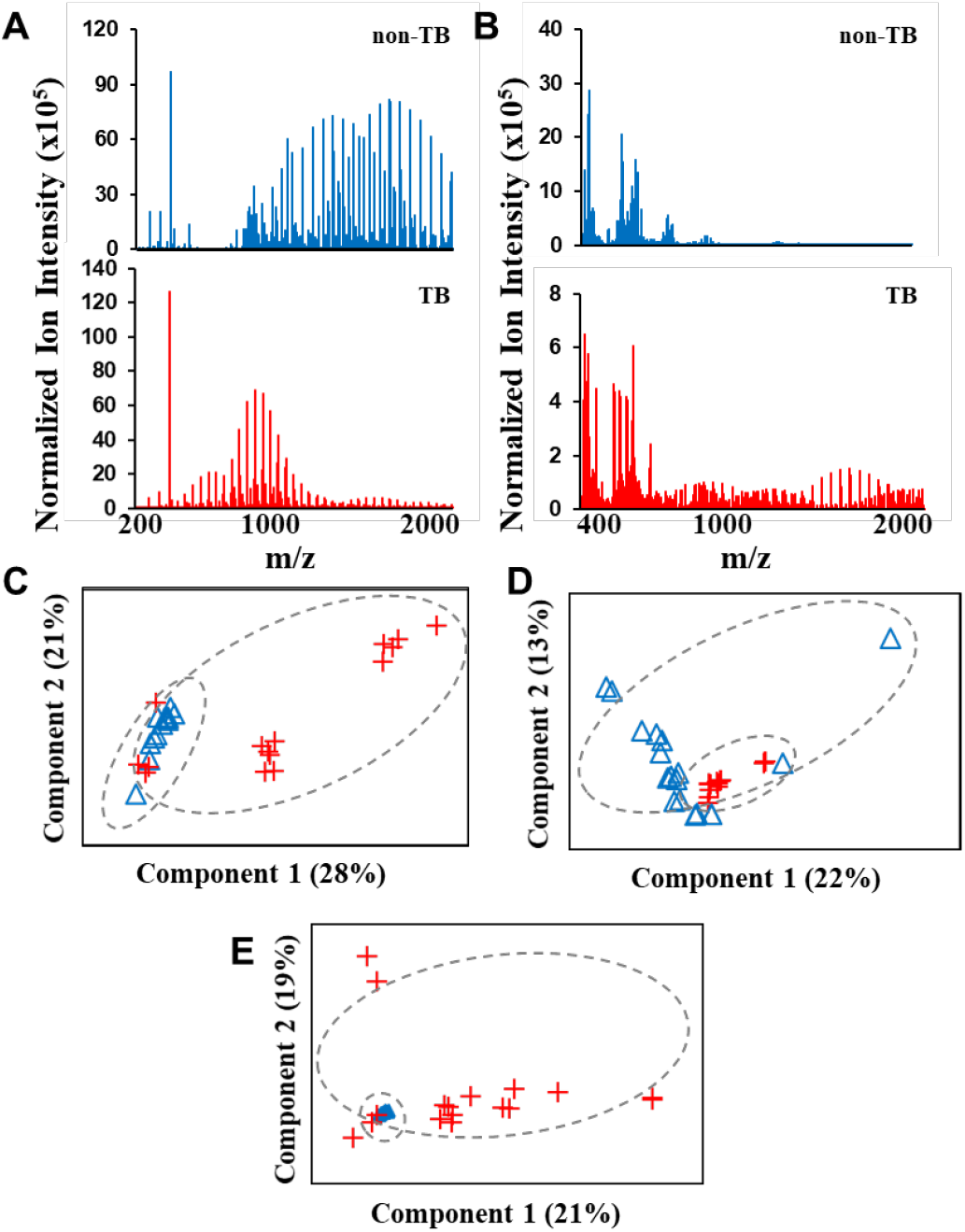
Representative high-resolution orbitrap mass spectra of breath samples and the analysis of mass spectrometric features by PCA. Data collection of active TB patients and non-TB subjects was collected in positive ion (**A**) and negative ion mode (**B**). Analysis was conducted with features collected from positive ion mode in the mass range of 200 to 2000 m/z (**C**) and negative ion mode in the mass range of 400 to 2000 m/z (**D**) and 900 to 2000 m/z (**E**).

### Significance analysis of microarray (SAM)-based feature selection

SAM was developed to select features with the strongest discriminative power to distinguish two classes of samples in omics studies^13^. In this study, SAM was adapted to visualize features that had the most powerful quantitative capacity to distinguish between non-TB subjects and TB patients. Generally, SAM returned a list of features ranked by statistical powers, fold-change, and false-positive rate^13^. SAM analysis showed that 1315 features were upregulated and 188 features were downregulated over the adjustable threshold (Table S2). Five features with the best scores were identified as 832.55 Da (C47 H79 O9 N P) with a fold-change of 443, 876.58 Da (C42 H87 O15 N P) with a fold-change of 455, 1114.66 Da (C61 H97 O15 N P) with a fold-change of 118, 744.50 Da (C36 H75 O12 N P) with a fold-change of 136, and 920.61 Da (C44 H91 O16 N P) with a fold change of 456 (Figs. 3 A-J, Table 1).

**Table 1.**
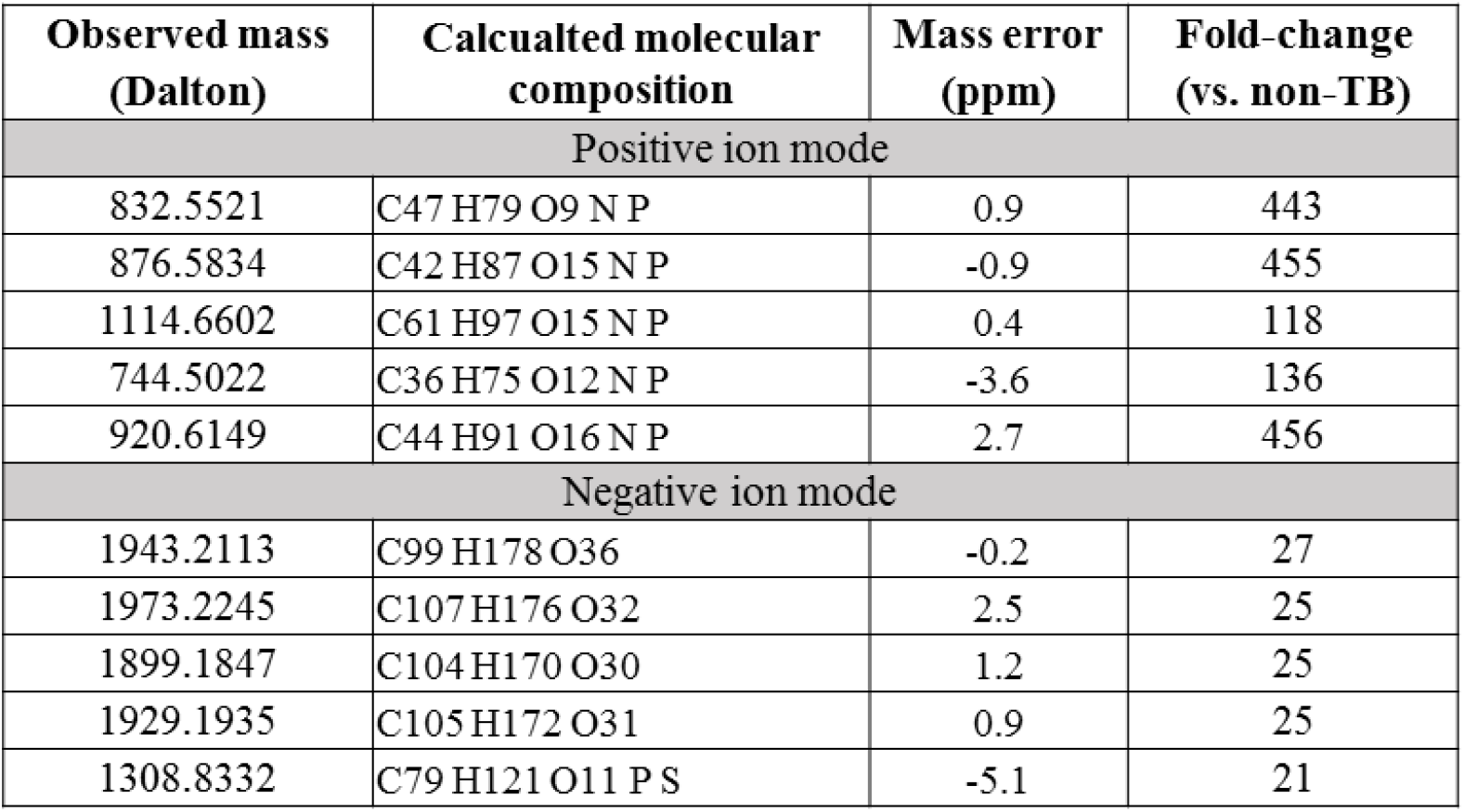
Top extracted features in positive ion and negative ion modes with SMA analysis.

**Figure 3.**
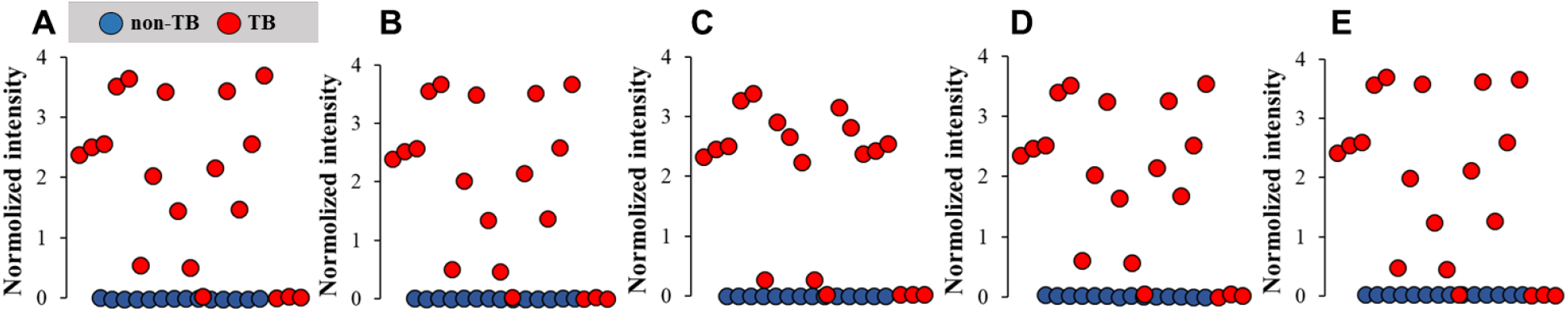
SAM analysis with extracted mass spectrometric signals collected in positive ion mode. The distribution of top-5 features that are significantly upregulated in non-TB subjects (blue solid dots) and TB patients (red solid dots) **(A-E)**. The fold-change distribution of top-5 features selected by SAM analysis **(F-J)**.

In negative ion mode, SAM analysis showed that 329 features were upregulated and 29 features were downregulated over the adjustable threshold (Figure 4 and Table S1). The best features with upregulated levels in TB patients were assigned to 1943.21 Da (C99 H178 O36) with a fold-change of 27, 1973. 22 Da (C107 H176 O32) with a fold-change of 25, 1899.18 Da (C104 H170 O30) with a fold-change of 25, 1929.19 Da (C105 H172 O31) with a fold-change of 25, 1308.83 Da (C79 H121 O11 P S) with a fold-change of 21 (Fig. 4, Table 1).

**Figure 4.**
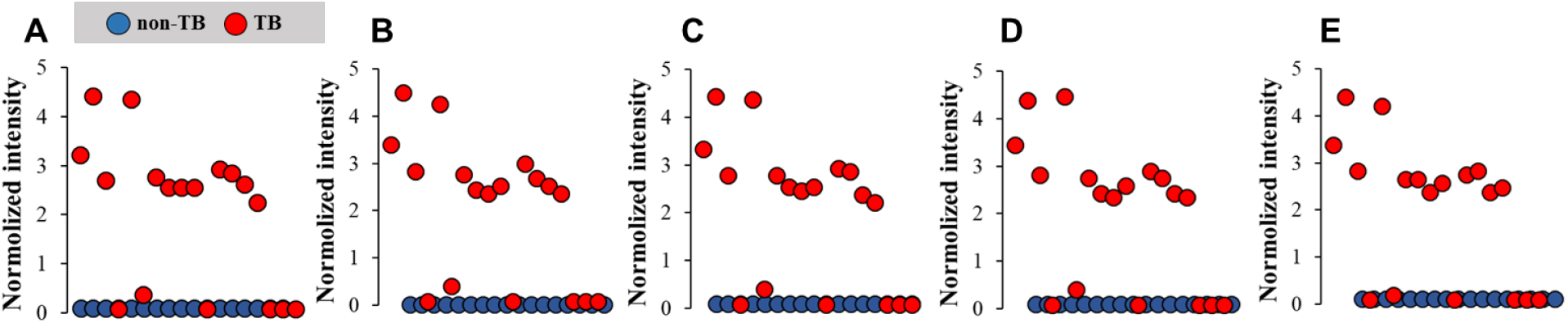
SAM analysis with extracted mass spectrometric signals collected in negative ion mode. The distribution of top-5 features that are significantly upregulated in non-TB subjects (blue solid dots) and TB patients (red solid dots) **(A-E)**. The fold-change distribution of top-5 features selected by SAM analysis **(F-J)**.

### Optimization of selected features using support vector machine (SVM)

In our study, over 4000 signals were routinely observed with high-resolution orbitrap mass spectrometry analysis from a small group of study participants (<30). Therefore, it is critical to construct a strategy to identify the most relevant TB biomarkers. By using the SAM-based feature selection method, over 2000 features were selected as the most powerful segregation factors. To further validate these selected features, SVM was introduced. The ideal situation for the application of SVM is to have both training and testing datasets. However, in our study we have limited numbers of study participants and SVM was, *per se*, used to evaluate the features prepared for the down-stream machine-learning work^14^.

The features with upregulated levels in TB patients were applied to SVM analysis and the ability to segregate two classes of samples were calculated to optimize the selected features. As expected, a result showed a low segregation percentage when all the features were applied, results from over-training. The best segregation percentage rate was observed when ∼ 300 features extracted from positive ion mode were applied (Fig. 5 A). For features extracted from negative ion mode, the best segregation percentage was observed when ∼ 100 features were applied (Fig. 5 B). Using this approach for positive ion mode data, we were able to distinguish actual 15 TB patients from non-TB subjects. Using negative ion mode data, we were able to reach the segregation percentage of 89%. Volcano plots were generated using the optimized features collected from positive ion and negative ion mode as another visualization tool to observe each feature based on the fold-change values and their statistical significance (Figs. 5, D and E). As expected, the optimized features with best scores with SAM analysis in each ion mode fall to the right corner of the plots which indicate higher quantitative values and statistical significance (red dots, Fig. 5). However, since volcano plots, *per se*, do not consider false-positive rates, the farthermost features in the plots do not necessarily provide the strongest distinguishing power. In addition, the volcano plots and SAM analysis revealed another important message showing that more features were upregulated in TB patients, which could be a result of detection of biomolecules shred from destructed lungs with modified pathology caused by TB infection.

**Figure 5.**
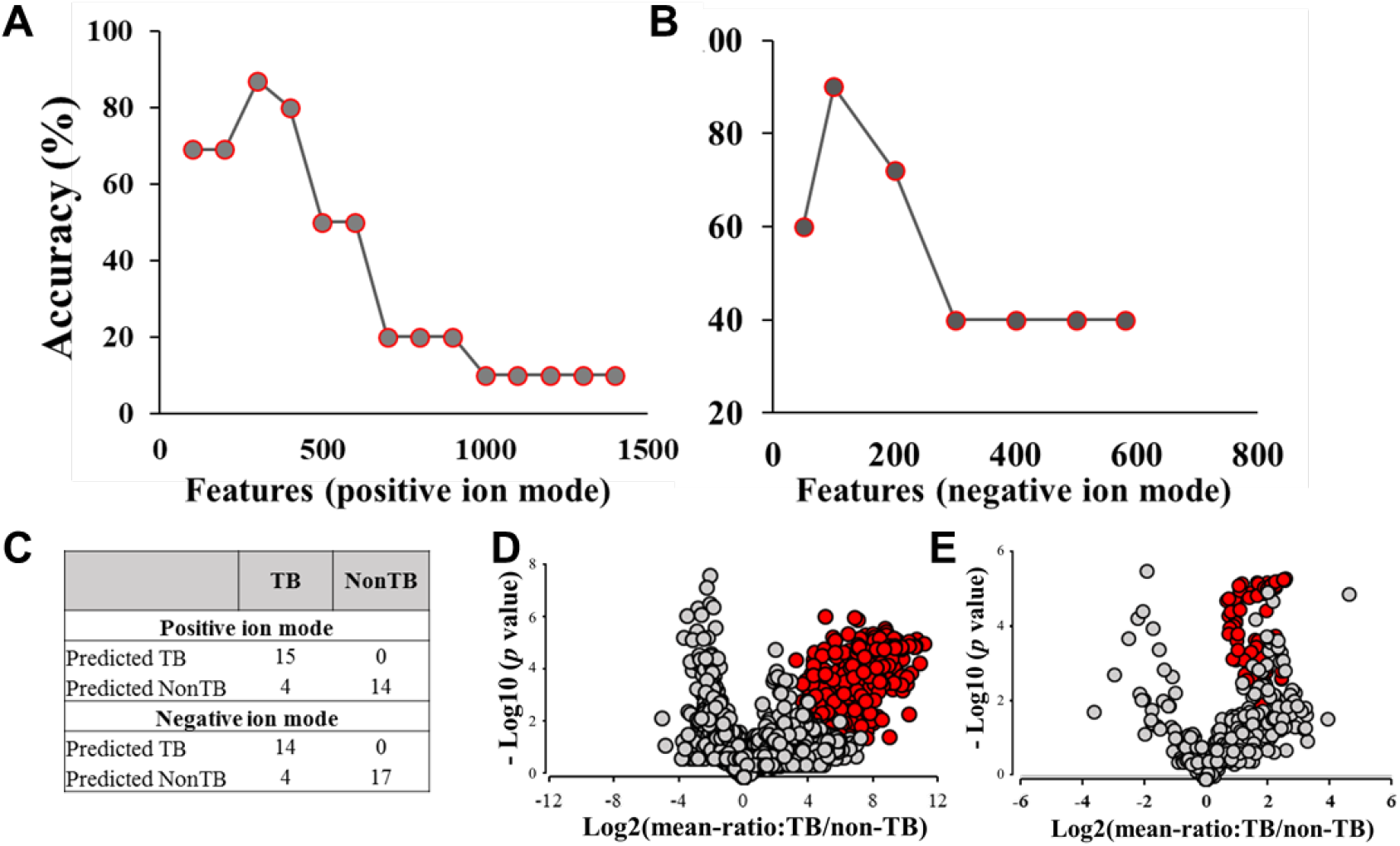
Optimization of selected features using support vector machine. Analysis was performed using selected features in positive ion (**A**) and negative ion mode (**B**) including an evaluation of segregation percentage between two classes of samples provided by optimal features (**C**). Red dots represent optimized features using SVM in positive ion **(D)** and negative ion **(E)** modes, respectively.

## Discussion

Human exhaled breath contains disease biomarkers and can be used as a noninvasive sample collection method for medical investigations^15^. A significant majority of breath analysis research focused on the measurement of volatile organic compounds (VOC) and several VOC detection-based tests were approved by the US Food and Drug Administration^16, 17^. Besides volatile compounds, exhaled breath air also contains nonvolatile compounds extracted from the respiratory tract lining fluid^18^. Since nonvolatile compounds are mostly identified as cytokines, phospholipids, and metabolites, it is highly likely that nonvolatile components contain more informative biomolecules associated with human disease states^19,20^. Many studies of the investigation of nonvolatile compounds have relied on the sample collection method exhaled breath condensate (EBC)^15^. In this technique, the subject’s exhaled air passes through a cooled object and forms condensed fluid^15^. A large number of candidate biomarkers can be identified for disease state using this strategy^21^. However, since there are no well-defined standards to quantify the analyte concentration, sample standardization is difficult using EBC collected samples^15^. In addition, saliva and sputum contamination during sample collection can add complications to the analysis^15^. These limitations contribute to the unreliability of measurement and low reproducibility using EBC collection methods.

The work we present here pushes beyond the EBC collection to capture particulate matter emanating from the lungs^22^. This exhaled breath aerosol (EBA) sample collection technique was developed to capture bioaerosol particles which contain lung surfactant and microparticle fractions associated with the transmission of infectious diseases^11,12^. EBA-based collection methods offer several advantages. Since EBA sampling methods focus on aerosol particles, study participants do not necessarily make direct contact with the sampling apparatus and saliva/sputum contamination can be avoided during the sample collection^11^. The respired aerosol particles are also analyzed using optical scatter spectrometers to provide a quantitative aerosol particle size distribution that enables sampling standardization^11,12^. In our previous study, we developed an aerosol sampling system, respiratory aerosol sampling chamber (RASC), for the isolation and collection of breath aerosol particles^11^. Preliminary studies conducting on newly diagnosed tuberculosis (TB) patients demonstrated that advanced EBA collection method had the capacity to isolate airborne particles and organic substances including *Mycobacterium tuberculosis*, as confirmed by microbiological culture and polymerase chain reaction method^11,12^. The success of RASC-based aerosol particle collection encouraged us to develop a strategy by which biomolecules contained in aerosol particles can be characterized and used for the detection of active TB cases.

Analysis of aerosol particles relies on various types of molecular-based technologies that usually deal with a small sample volume such as mass spectrometry^23^. The final volume of aerosol particle sample is ∼ 10 mL of buffer solution, meaning it requires an enrichment step. For this purpose, we employed Folch extraction, one of the most commonly used methods for lipid extraction^24^. Advantages of this enrichment method rely on the nature of our breath samples. It was reported that the major components in aerosol particles are lung surfactant that contains phospholipids and *Mycobacterium* nonvolatile molecules^11,12,22^. Those analytes can be feasibly introduced to mass spectrometry by conventional ionization methods. In addition, the molecular mass of common phospholipids is from 200 to 2000 Da, which matches excellently with the capacity of the high-resolution orbitrap mass spectrometer. Moreover, for lipid analysis, signal collection can be conducted in both positive ion and negative ion modes^25^. The combination of both data collection methods enables a deep mass spectrometry analysis and increase opportunities for detecting potential TB biomarkers. During lipid extraction, the aqueous phase, which contains mass spectrometry-interfering salts and chemicals, is isolated and excluded from the final sample. In this way, the enrichment method serves as an essential step for sample clean-up and desalting for sequential mass spectrometry analysis.

Mass spectrometry has been widely used for the characterization of breath samples and identification of biomarker candidates^26^. Those biomarkers include protein cytokines, surfactant lipids, and metabolites for various human diseases such as for breast cancer, chronic obstructive pulmonary disease (COPD), and asthma^12,28,29^. During untargeted mass spectrometry-based profiling for potential biomarker identification, chromatographic separation is commonly used to achieve a deep characterization of biological samples^30^. However, the chromatographic step is time-consuming and cannot be applied to fast screening when large populations and repeated testing is involved^30^. The high-performance mass resolving capacity of the orbitrap system enables chromatograph-free direction infusion mass spectrometry (DIMS)^31^. To evaluate whether the signals collected from DIMS could reveal essential differences between non-TB subjects and TB patients, we performed an unsupervised principal component analysis on signal profiles of each study participant without prior knowledge of patient disease state. The results showed that segregation between two classes of samples was present. Follow up unblinded analysis showed that TB patients clustered nicely with separation from the non-TB patients. However, several TB patients were prone to cluster with non-TB subjects in both ion modes. Interestingly, study participant information showed that they belong to the same TB patients, suggesting that examination of the metadata associated with patients’ age, gender, sputum analysis, chest X-ray, HIV status, history of previous TB disease, cough rate, etc, should be considered to reveal this disparity. In addition, PCA results of negative ion mode showed better segregation between non-TB and TB samples in the mass range of 900 to 2000 Da. This observation agreed with our mass spectrometry analysis in which most signals generated below 900 Da were visually identical and distinguishable peaks were observed in the mass range of 900 to 1000 Da. Although unsupervised PCA and cluster analysis provides coherent profile patterns between non-TB subjects and TB patients, statistical significance is required to identify TB-relevant features. In addition, thousands of signals were collected from relatively small numbers of study participants and methods are required to evaluate if signal profiles are experimentally significant. To address these challenges, we developed a strategy combining a feature selection method, significance-analysis of microarray (SAM), and a machine-learning algorithm, support vector machine (SVM), to extract the most discriminative features that can distinguish non-TB subjects and TB patients^13,14^. The principle of SAM is to generate a ranking of features according to a score assigned to each feature based on statistical significance, relative abundance, and false-positive rate. Then, the selected features were trained in SVM as predictors that can provide as much as segregation between non-TB subjects and TB patients. The advantages of this strategy rely on its ability to reduce the complexity of the high-dimensional dataset, identify as many as features with great statistical power, and optimize features to avoid overfitting. In addition, this data processing strategy has the potential to be applied to lipid species that were well identified from *Mycobacterium tuberculosis* to achieve a more comprehensive analysis for TB biomarkers by using the recently developed *Mtblipid* database^32,33^ and lipidomics analysis of *Mycobacterium* tuberculosis^34^.

Chemical formula analysis using accurate mass measurement-based composition prediction revealed that the features with the most statistical power belonged to phospholipid classes that were strongly induced in TB patients. Previous studies showed that phospholipids, such as dipalmitoylphosphatidylcholine (DPPC), were the main components of human pulmonary surfactant^35^. This is consistent with the observations that TB patients suffer from damaged lung due to *Mycobacterium tuberculosis* infection. Alternatively, since individual particles of *Mycobacterium tuberculosis* has been observed in the aerosol contents using the RASC^11,12^. Therefore, we cannot rule out the possibility that phospholipids had the origin from *Mycobacterium tuberculosis* bacterial wall. Indeed, features with the most statistical power in negative ion mode collection were predicted as glycolipids, which were recently found to be highly abundant in the mycobacterial cell envelope. Our observations prompt the speculation that the biomolecules contained in aerosol particles collected in the RASC could be from both cellular components of host lung and the mycobacterial envelope. However, the exact molecular composition of identified features in this study needs to be confirmed by the interpretation of fragmentation patterns offered by tandem mass spectrometry.

In this study, we developed a strategy in which direct infusion was applied to sample introduction and the characterization of molecular formula relied on accurate mass measurement-based composition prediction. By using this strategy, signals collected from both positive and negative ion were inclusively characterized. Additionally, since no tedious separation step is needed, this strategy holds the potential to be developed to a fully automated and fast analytical method for the rapid identification of TB in heavily affected areas. For this aim, we plan to collect an additional amount of breath samples from both healthy subjects and TB patients. In this way, mass spectrometric datasets can be used to train our model better and evaluate the features extracted from our machine-learning algorithms. In addition, for sample preparation, although Folch extraction-based enrichment method is classic and used extensively, it is time-consuming. Notably, several rapid lipid extraction methods, such as solid-phase extraction (SPE), have been developed and will be tested in future studies^36^. Another advantage of using SPE-based extraction is that proteins contained in the aerosol particles can be preserved. Mass spectrometry-based rapid identification of proteins without enrichment and enzymatic digestion were developed^37^. As a result, signature proteins characterized by TB patients could be combined with lipid biomarkers to provide a more reliable diagnostic approach for the identification of active TB patients.

## Materials and Methods

### Statement and patient information

All participants provided written informed consent and the study was approved by the University of Cape Town Human Research Ethics Committee (Reference number HREC 680/2013). All methods were performed in accordance with relevant guidelines and regulations. Exhaled breath samples were collected from 19 TB patients and 17 non-TB subjects at the Desmond Tutu HIV Centre near Cape Town (South Africa) from August 2017 to February 2018. The diagnosis of active TB was based on a positive sputum GeneXpert™. HIV and smoking status are demonstrated in Table S1.

### Exhaled breath aerosol particle collection

Exhaled breath particle collection was conducted in the respiratory aerosol sample chamber (RASC) unit reported in previous studies (Wood et al., 2016; Patterson et al., 2017). Aerosol-only samples were collected from study participants as they breathe, and spontaneously cough, in the RASC unit for 60 minutes. The bio-aerosol particles were continuously collected using a 240 liter *per* minute wetted well cyclone collector (Coriolis *μ* Biological Air Sampler, Bertin Instruments, Montigny-le-Bretonneux, France) into 10 mL of phosphatebuffered saline solution.

### Sample concentration with lipid extraction

All chemicals are MS-grade and acquired from Fisher Scientific (Thermo Fisher Scientific, MA, USA). Lipid standards were acquired from Matreya, LLC (State College, PA). 5 mL of stock liquid breath sample was moved into a 15 mL conical tube and lyophilized overnight. After lyophilization, freeze-dried samples were mixed with 1 mL of chloroform: methanol (2:1, v/v) solution and left to sit at room temperature for 1 h. Then, 250 *μ* L of MS-grade water was added, vortexed, and left to sit at room temperature for 10 min. The samples were then carefully moved to a microcentrifuge and phase separation was finished by spinning at the speed of 5,000 × *g* for 10 min. The lower phase samples, which contain lipid molecules, were carefully collected and dried with dry nitrogen gas at room temperature. The recovered samples were reconstituted in 100 *μ* L of chloroform: methanol (2:1, v/v) solution. Prior to mass spectrometry analysis, 10 *μ* L of samples were diluted with 90 *μ* L of acetonitrile: isopropanol: water (2:1:1, v/v/v/) and 0.1% formic acid. Two lipid standards were processed using the same protocol as quality control.

### High-resolution mass spectrometry analysis

Samples were introduced to an LTQ orbitrap mass spectrometer (Thermo Fisher Scientific) via electrospray ionization by direct infusion (flow rate: 3 *μ* L/min). Positive and negative ion calibration solutions (Thermo Fisher Scientific) were used for ion source optimization and mass accuracy test. For initial data acquisition, a resolving power of 60, 000 at 200 m/z was used for both positive and negative ions in the mass range of 200 – 2000 m/z. For positive ion mode data collection, we included 19 TB subjects and 14 non-TB subjects. For negative ion mode data collection, we included 18 TB subjects and 17 non0TB subjects.

### Mass spectrometric signal processing and data analysis

Raw data was processed on Xcalibur 3.0 Qual Browser (Thermo Fisher Scientific). For deconvolution and peak isolation, mass peaks and intensity values were extracted with a signal to noise ratio over 5 with 60,000 resolution. Elemental compositions for each extracted mass peak were calculated using Xcalibur 3.0 Qual Browser with a mass tolerance of 5 ppm: element options as 150 carbon (C), 300 hydrogen (H), 100 oxygen (O), 1 phosphorus (P), 1 sulfur (S), and 1 nitrogen (N). Ion intensity values were used for data normalization. For normalization, the monoisotopic peaks were deconvoluted using Xcalibur 3.0 Qual Brower software. The peak list generated from the deconvolution process includes masses, resolution values, molecular compositions, and individual peak’s relative ion intensity by the percentage (the most intense peak is labeled as 100%). The molecular composition and its corresponding percentage values were extracted and used for data analysis. Principal component analysis (PCA) was applied to normalized data for the visualization of the segregation between non-TB subjects and TB patients. For feature selection, significance analysis of microarray (SAM) was performed using RStudio. SAM is a well-developed tool to serve omics studies and returns a list of features ranked by arbitrary scores based on their statistical power, fold-change, and false-positive rate (Tusher et al., 2001). A machine-learning algorithm, support vector machine (SVM), was used to optimize the features selected by SAM (Suykens et al., 1999). SVM reduces high dimensional data, defines separation hyperplane, and segregates two classes of samples as clearly as possible (Suykens et al., 1999).

The performance of feature selection was presented segregation percentage (total number of two correct predictions divided by the total number of a dataset) as the ability to segregate TB and non-TB subjects.

## Data Availability

The authors confirm that the data supporting the findings of this study are available within the article and its supplementary materials.

## Acknowledgments

We thank Dr. Catherine Fenselau (Professor Emeritus, Dept. of Chemistry & Biochemistry, University of Maryland, 2018-present) for technical and analytical expertise. RW was partially funded by the Strategic Health Innovation Partnership (SHIP) Unit of the South African Medical Research Council, as a sub-grant from the Bill and Melinda Gates Foundation.

## Author contributions

W.A.B and R.H. collected breath samples. D.C. and W.A.B contributed to the mass spectrometry research design. D.C. performed mass spectrometry analysis, data processing, and statistics. D.C. drafted the main manuscript and R.H. prepared the methods for breath sample collection. All authors understood the data interpretation and agreed upon the results. All authors reviewed and approved the manuscript.

## Supplementary materials

Fig. S1. High-resolution mass spectrometry analysis of lipid standards.

Table S1. Active TB patient and non-TB subject information.

Table S2. Raw data files of SAM analysis of features extracted from positive ion and negative ion mode.

